# A comparison of the EQ-5D-5L and the SF-6Dv2 among patients with hypertrophic obstructive cardiomyopathy

**DOI:** 10.1101/2023.10.25.23297528

**Authors:** Xiaoyu Wang, Haiqiang Sang, Rui Meng, Xin Su, Peng Liu

## Abstract

**Background:** Hypertrophic obstructive cardiomyopathy (HOCM) is a serious and complex chronic disease severely affecting patients’ daily lives and health-related quality of life (HRQoL). The psychometric performance of generic preference-based instruments has not been compared in Chinese patients with HOCM. We aimed to identify an applicable vehicle to determine HRQoL and explore the psychometric properties of SF-6Dv2 and EQ- 5D-5L in adults with HOCM. The interchangeability of the tools in cost-utility analysis was also investigated.

**Methods:** We collected data from 131 patients with HOCM from the First Affiliated Hospital of Zhengzhou University, China. Assessments were performed on the day of admission and three and six months after discharge using SF-6Dv2 and EQ-5D-5L. The responses were converted to utility values using the corresponding Chinese value sets. The tool distributions were explored, and the floor and ceiling effects were analyzed. The agreement was assessed using intra-class correlation coefficients (ICCs) and Bland–Altman plots. Convergent validity was tested using Spearman’s rank correlation coefficient. Known group validity was measured across various clinical and sociodemographic indicators using relative efficiency (RE) statistics.

**Results:** The mean utility scores for SF-6Dv2 and EQ-5D- 5L at baseline and 6-month follow-up were 0.61, 0.62, 0.736, and 0.797, respectively. The EQ-5D-5L and SF-6Dv2 distribution scores showed no normality. EQ-5D-5L was more sensitive to changes over time and showed a moderate to good correlation with the SF-6Dv2 (ICC: 0.598–0.862). The instruments’ agreement and convergent validity worsened in patients with a higher New York Heart Association (NYHA) cardiac function classification and lower general health status. SF-6Dv2 showed higher relative efficiency statistics and a greater ability to distinguish external health status.

**Conclusions:** The measured results can be used for future cost-utility analyses. SF-6Dv2 and EQ-5D-5L presented different results and should not be used interchangeably. SF-6Dv2 is optimal for detecting differences between subgroups with various health states.

## Introduction

Hypertrophic cardiomyopathy (HCM), distinguished by left ventricular myocardial hypertrophy, is a class of cardiac disorders caused by genetic factors. It often has an autosomal inheritance pattern due to variants in the genes encoding myosin (or myosin-associated genes) or may have an unknown genetic etiology [1]. The main feature of hypertrophic obstructive cardiomyopathy (HOCM) is a left ventricular outflow tract pressure difference of at least 30 mmHg at rest or with provocation. It accounts for approximately 70% of all HCM patients [2]. Currently, the prevalence of HCM in China is 0.076%, and the mortality rate of HCM patients is as high as 3.38% [3]. HOCM is the main cause of sudden cardiac death in adolescents and athletes and is closely related to heart failure and stroke in older patients [4,5]. Typical symptoms include exertional dyspnea and weakness, chest pain, and palpitations, which tremendously impact patients’ physical health and health-related quality of life (HRQoL) [6]. Parallel to this, HOCM is closely related to considerable expenditures on healthcare and imposes an enormous financial burden on global health budgets [7].

In order to provide evidence support for decision-makers to allocate limited resources among competing healthcare programs appropriately, cost-utility analysis (CUA) is incrementally being used. Quality-adjusted life years (QALYs) are health outcome indicators in CUA, which combines years of survival with utility values [8]. Utility scores are usually expressed as a value from 0 to 1, where 1 represents perfect health and 0 represents death; they can also be negative, reflecting a disease situation worse than death. A series of universal health scales (multi-attribute utility instruments [MAUIs]) have been developed internationally to measure health utility values. Among the multitude of generic preference-based scales, the EuroQoL Five-Dimension (EQ-5D) from the European Society of Quality of Life [EuroQoL Group] along with the Short Form Six-Dimension (SF-6D) obtained based on the Short Form 36-item [SF-36] are among the most popular [9].

EQ-5D, developed by the EuroQol Group, is the most commonly used MAUIs worldwide. Compared to the original version of EQ-5D-3L, which contains only three response levels for each dimension, EQ-5D-5L offers five levels per dimension and lower ceiling effects when conducting a survey among the population [10]. The SF-6D scale is second only to the EQ-5D. Moreover, because the SF-6D scale dimension level setting is more abundant and sensitive, it is more suitable for chronic diseases where the clinical symptoms are not obvious. Compared with SF-6D, SF-6Dv2 has a much broader scoring range and addresses the problem of descriptive systems [11]. Differences in the structure and valuation of different instruments may give rise to various estimates of the identical “health state” for the same individual, which results in discrepancies in utility and healthcare-related decision-making. Therefore, it is crucial to measure the utility of HOCM patients using these two well-known instruments and to compare the psychometric properties of measurements and interchangeability.

Comparisons of the properties of SF-6D and EQ-5D measurements have already been validated for several diseases, including depressive symptoms and fibromyalgia [12,13].

However, no studies have compared health utility values based on Chinese HOCM patients using EQ-5D-5L and SF-6Dv2. Therefore, we aimed to estimate the utility of Chinese patients with HOCM using EQ-5D-5L and SF-6Dv2. Furthermore, we evaluated whether EQ-5D-5L and SF-6Dv2 revealed similar empirical estimates of health-related utility and interchangeability in patients with HOCM.

## Material and methods

### EQ-5D-5L

The EQ-5D-5L consists of two parts: a questionnaire section and the EuroQoL-visual analog scale (EQ-VAS). The questionnaire contained five dimensions, each with five severity levels: no problem, mild, moderate, severe, and very severe [14]. Thus, 3125 (5 × 5 × 5 × 5 × 5 × 5) different health states described by the EQ-5D-5L questionnaire can be calculated, of which 11111 are the best health states (perfectly healthy) and 55555 are the worst state of health. The EQ-5D-5L utility value integral system converts a patient’s health status into health utility value. A Time-trade off (TTO) approach was employed to develop Chinese EQ- 5D-5L utility values ranging from –0.391(55555) to 1(11111). Furthermore, the EQ-VAS indicates the patient’s subjective rating of self-reported health state on the day being interviewed, with a vertical length of 20 cm, starting at 100 (the best health imaginable) and ending at 0 (the worst health imaginable) [15]. The value of the EQ-VAS was converted to 0- 1 to facilitate comparison.

### SF-6Dv2

The SF-6Dv2 serves a broader range of dimensions than the EQ-5D-5L, of which only ten items in the SF-36 have been reclassified into six items, each dimension corresponding to one item [16]. All the dimensions except pain domains (six levels) had five levels, yielding a total of 18,750 (= 5 × 5 × 5 × 6 × 5 × 5 × 5) health conditions [17]. We use the set of Chinese tariff values developed for SF-6Dv2 by the TTO method, which has a range of values from -0.277 (555,655) to 1 (111,111).

### Sampling and data collection

Participants were patients with HOCM who were hospitalized at the First Affiliated Hospital of Zhengzhou University between 1 June 2021 and 31 October 2022. Patient demographic characteristics and disease-related data, including age, sex, work and marital status, educational background, body mass index (BMI), and left ventricular ejection fraction (LVEF), were completed by the patient or extracted from the inpatient electronic medical records.

The inclusion criteria were supposed to be in accordance with the following: 1) consistent with the diagnosis of hypertrophic obstructive cardiomyopathy, and 2) the completed scale contained no missing data. Patients were excluded if they 1) were under 18 years old, 2) refused to sign the informed consent form, or 3) had other serious illnesses or psychological disorders that prevented them from understanding the contents of the questionnaire. Eligible respondents were interviewed face-to-face on the day of admission by a research assistant who underwent a 1-day uniform professional training before the study started. After filling in the questionnaire, a second research assistant verified that the questionnaire was complete and that no items were missing. Following discharge, the patient was contacted by phone three and six months later. Upon completion of the questionnaire, the patients uploaded and returned the results.

The study protocol was approved by the University of Zhengzhou Ethics Committee (registration number 2021-KY-0943-002), and all participants provided written informed consent before participating in the research. This study adhered to the tenets of the 1975 Declaration of Helsinki (Tokyo revision, 2004).

### Statistics analysis

The baseline demographic statistics described continuous and categorical variables using mean, standard deviation, and frequency, respectively. Overall quality of life (QOL) indices were calculated and compared for the SF-6Dv2 and EQ-5D-5L questionnaires at three time points: on the day of admission and three and six months after discharge. A histogram represented the distributions of the two instruments. Normality tests were conducted using the skewness and kurtosis values. Because of the highly skewed distribution of the index scores, the Wilcoxon signed-rank test for differences between the index scores of the two measurements was used. In contrast, the Kruskal–Wallis test was used for comparisons between the index scores of the participants’ characteristic groups. One-way ANOVA was applied to measure changes in New York Heart Association (NYHA) cardiac function classes during follow-up. Ceiling and floor effects were measured by computing the percentage of respondents who received the highest or lowest scores in each field. The minimally important difference (MID) is often applied to explain patient-approved minimum clinical effectiveness questionnaire score changes reflecting HRQoL. The treatment is clinically significant when the score change reaches the MID threshold. In a previous study, the average MID values of the SF-6Dv2 and EQ-5D-5L were 0.041 and 0.074, respectively [18]. Intraclass correlation coefficients (ICCs) were calculated for the SF-6Dv2 and EQ-5D-5L instruments to assess agreement, which was reported as a two-way random and absolute agreement with a single metric model: ICC lies within 0.5-0.75 for moderate, 0.75-0.9 for good and more than 0.9 for excellent. Bland–Altman plots were used to explore the degree of consistency between the two utility scores [19]. Convergent validity is the degree to which the results correlate when different measures are utilized to measure similar characteristics, such as anxiety/depression on the EQ-5D-5L and mental health on the SF-6Dv2. Convergent validity was evaluated by calculating Spearman’s correlation (ρ) coefficient between the domains of the two instruments. The strength of the Spearman correlation was rated as weak for less than 0.3, strong for more than 0.5, and moderate for in-between [20]. Known-group validity refers to the ability of two instruments to distinguish between each respondent’s characteristic group by calculating the average utility value for each measure and comparing them [21]. We examined how well SF-6Dv2 and EQ-5D-5L detected differences in the indices of external health based on the relative efficiency (RE) statistic. The RE is defined as the ratio of the square of the t-statistic of the comparison tool to the reference tool [22]. A value > 1.0 represents, when determining differences in health-related external indicators, EQ-5D-5L performs better than SF-6Dv2, while a value < 1.0 illustrates that the EQ-5D-5L performs less efficiently than SF-6Dv2.

Statistical analyses were performed using the SPSS, IBM Statistics software, version 22.0. In all statistical tests, p-values less than 0.01 were assumed to be statistically significant.

## Results

### Characteristics of the study sample

Four of the 135 participants died during the follow-up period. As their questionnaires were incomplete, they were excluded from the sample. Consequently, 131 patients with HOCM were included in this study. Participants’ sociodemographic and clinical characteristics at baseline are displayed in Tables 1 and 2, who were 53.4 years old, and most patients with HOCM were male, accounting for 55.7%, and 91.6% of patients were married. The mean body mass index (BMI) was 25.8 kg/m^2^. NYHA functional class III was present in 53.4% of the patients, and the LVEF ranged from 29% to 79%, with an average of 64.4%.

**Table 1.** Baseline characteristics.

| Characteristics | Total (n=131) |
| --- | --- |
| Age, mean (SD) | 53.4 (12.1) |
| BMI, mean (SD) | 25.8 (3.2) |
| EF%, mean (SD) | 64.4(5.66) |
| EQ-5D-5L utility, mean (SD) | 0.62(0.28) |
| SF-6Dv2 utility, mean (SD) | 0.61(0.19) |
| EQ-VAS score, mean (SD) | 0.62(0.18) |

SD: Standard deviation BMI: Body Mass Index; EF: Ejection Fraction; EQ-5D-5L: 5-level EuroQol 5-dimension; SF-6Dv2: Short Form Six-Dimension (version 2); EQ-VAS: EuroQol-Visual Analog Scale (EQ-VAS)

**Table 2.**
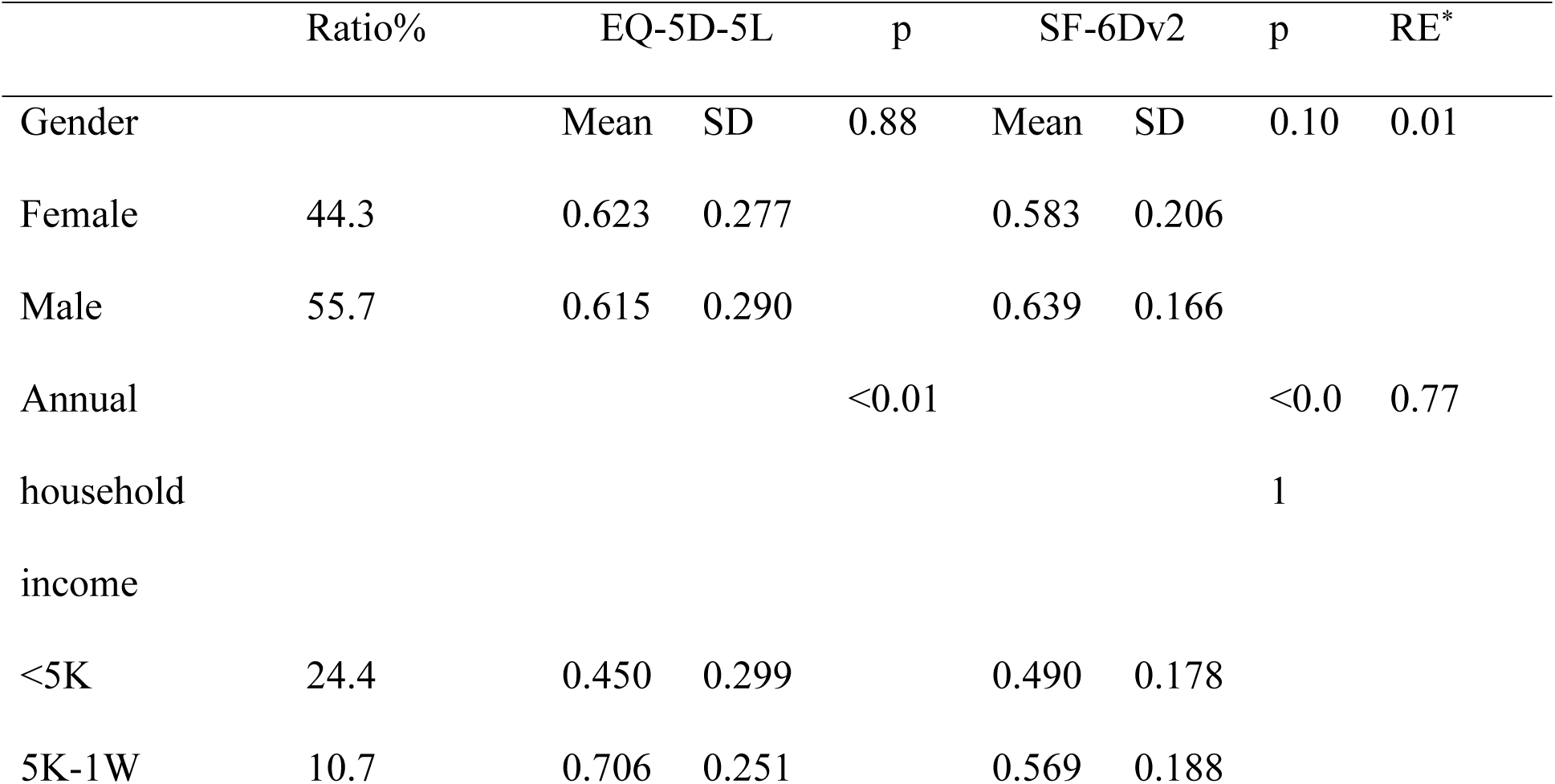
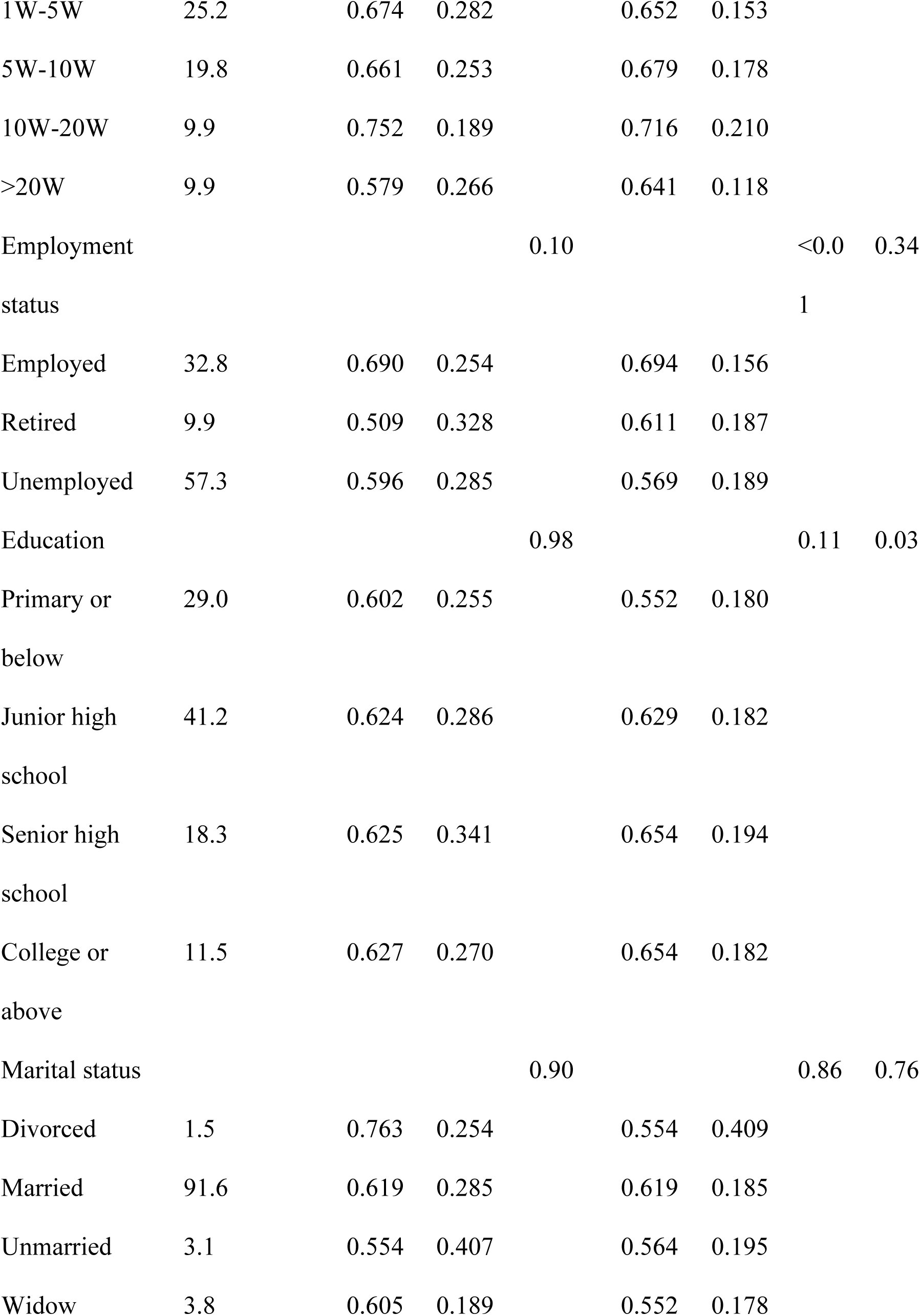
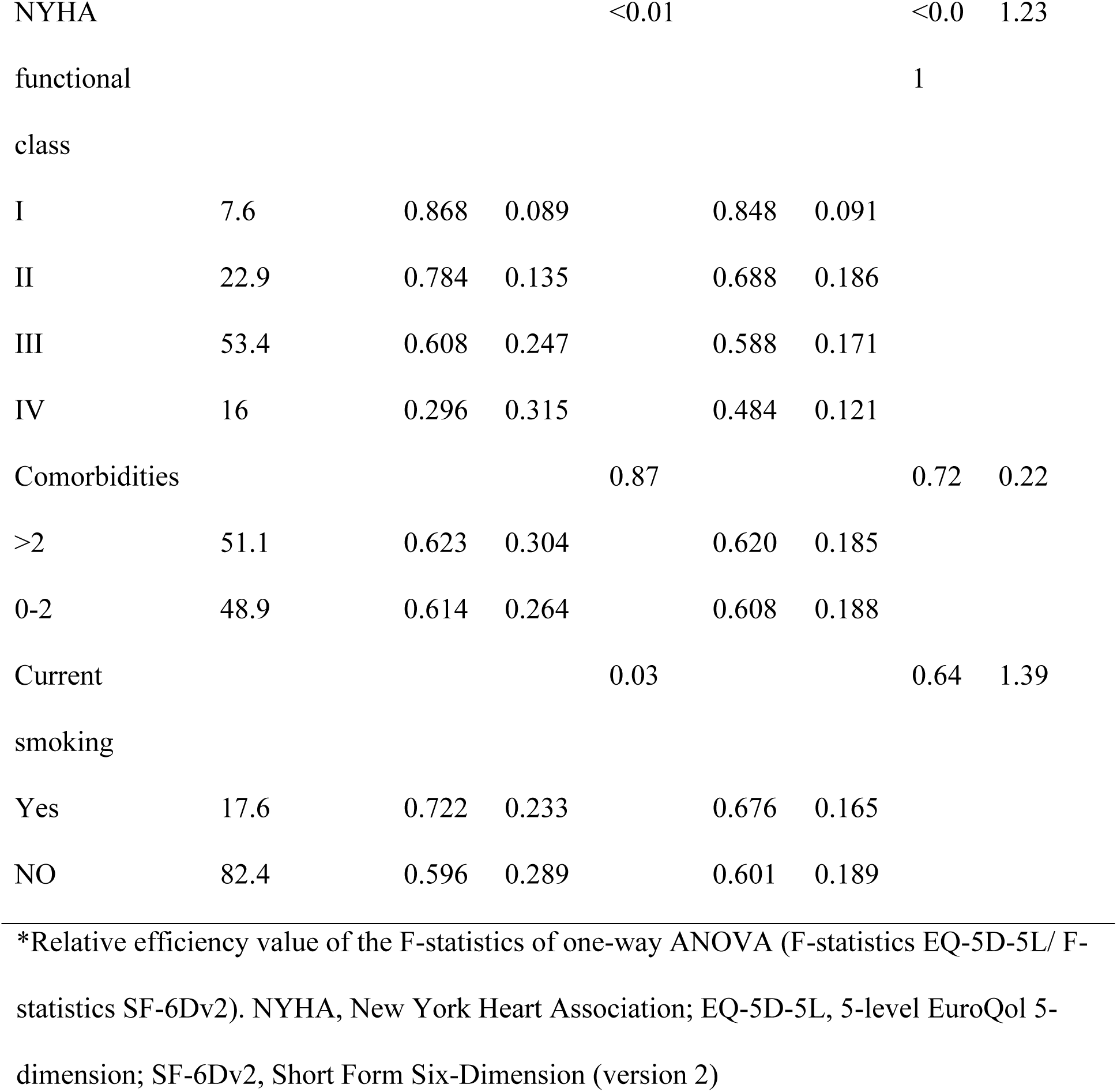
Known-groups validity of SF-6Dv2 and EQ-5D-5L.

We observed statistically significant differences in the NYHA cardiac function classification on the day of admission and at three and six months after discharge (p<0.01).

### Comparison

As shown in Fig 1, the scores from the baseline and follow-up measurements were not normally distributed. The distribution of EQ-5D-5L as a starting point and follow-up was highly left-skewed compared to SF-6Dv2. The most frequently distributed combinations of SF-6Dv2 and EQ-5D-5L at baseline were 421111 and 11121. The mean utility scores of SF- 6Dv2 and EQ-5D-5L at baseline were 0.61 and 0.62, respectively. The minimum and maximum values generated from the EQ-5D-5L were -0.182 and 1.0, respectively, while the

**Fig 1.**
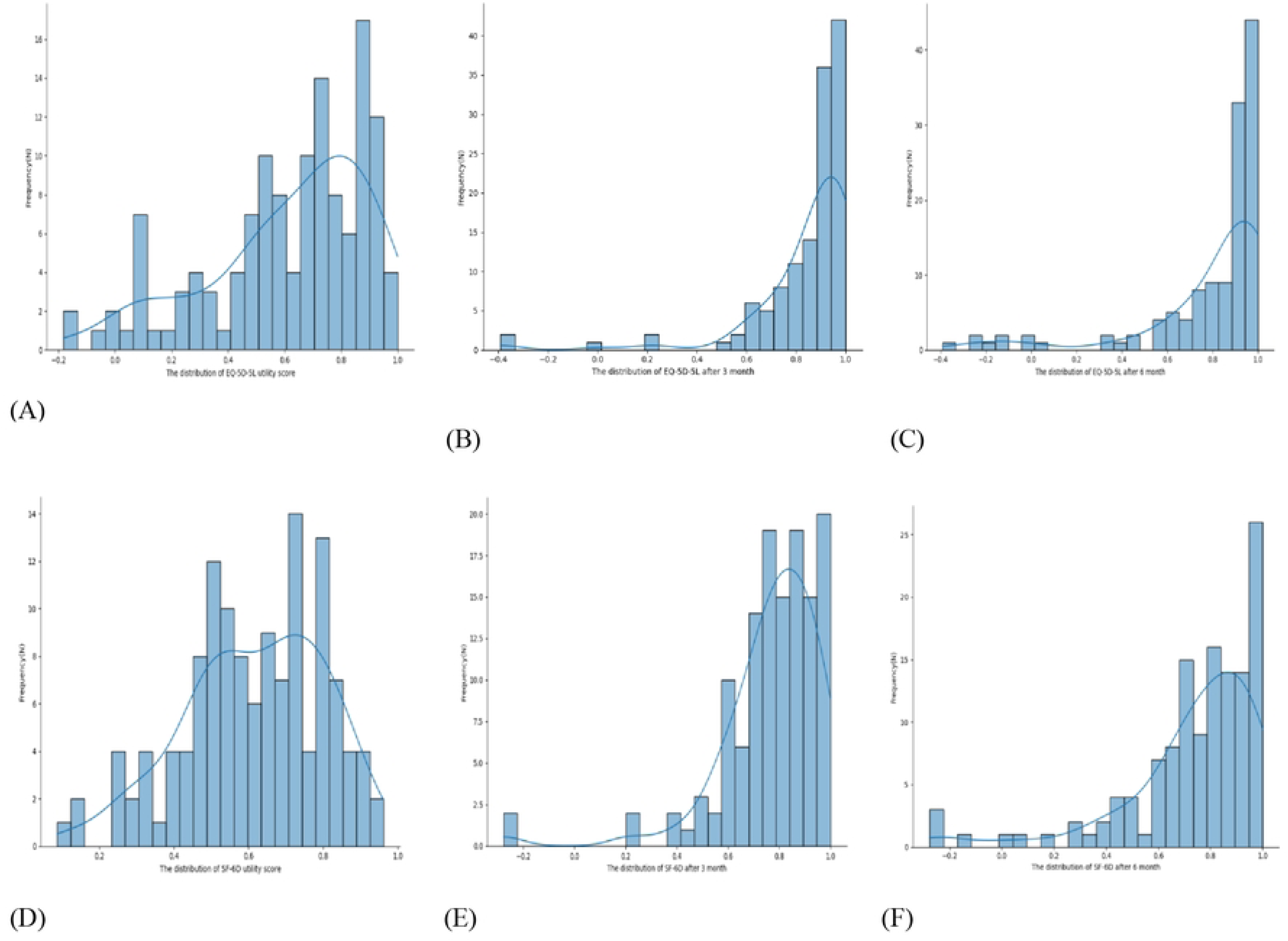
Distribution of EQ-5D-5L and SF-6Dv2. (A) Distribution of EQ-5D-5L at baseline; (B) Distribution of EQ-5D-5L at 3-month follow-up; (C) Distribution of EQ-5D-5L at 6- month follow-up; (D) Distribution of EQ-5D-5L at baseline; (E) Distribution of EQ-5D-5L at 3-month follow-up; (F) Distribution of EQ-5D-5L at the 6-month follow-up. EQ-5D-5L: 5- level EuroQol 5-dimension. SF-6Dv2: Short-Form Six-Dimensions (Version 2)

SF-6Dv2 measured –0.277 and 0.962 at its minimum and maximum, respectively. In EQ-5D- 5L, a low ceiling effect of 3 (2.3%) was observed. The EQ-VAS measured two (1.5%) participants in optimum health. No instruments produced a floor effect. Over a 3-month follow-up, the mean utility of EQ-5D-5L and SF-6Dv2 was 0.849 (range: –0.391 to 1.0) and 0.769 (range: –0.277 to 1.0), respectively (p = 0.000). The floor and ceiling effects of EQ- 5D-5L were 2 (1.5%) and 36 (27.5%), respectively, while those of SF-6Dv2 were 2 (1.5%) and 9 (6.9%), respectively. According to the EQ-VAS score, eleven individuals (8.4 %) obtained the best possible health. The 6-month follow-up revealed the following: the mean utility values of EQ-5D-5L and SF-6Dv2 were 0.797 and 0.736 (p = 0.000), respectively. The range of values of the two measurements was the same as for the follow-up in March. The floor and ceiling effects of EQ-5D-5L were 1 (0.8%) and 35 (26.7%), respectively, whereas those of SF-6Dv2 were 1 (0.8%) and 8 (6.1%), respectively; and 14 (10.7%) people achieved the highest EQ-VAS scores.

### Agreement

The overall agreement for the SF-6Dv2 and EQ-5D-5L in utility scores at baseline was moderate (ICC = 0.598). The ICCs at the 3- and 6-month follow-ups indicate good agreement when comparing EQ-5D-5L and SF-6Dv2 index scores (S1 Table). The Bland– Altman plots in Fig 2a show an average difference of 0.004, with a wide range of agreement from –0. 418 to 0.427 between the ED-5D-5L and SF-6Dv2 index scores. Consequently, the EQ-5D-5L measurement was 35, which was 9% less or 58% larger than the measurement by SF-6Dv2 for 95% of individuals. SF-6Dv2 and EQ-5D-5L had a large discrepancy for lower utility values, with high variation between the two instruments depending on the patient’s health status. For better health, EQ-5D-5L produced higher scores, while for poorer health, SF-6Dv2 yielded higher scores. The 3-month follow-up period is shown in Fig 2b. The indicator scores for EQ-5D-5L exceeded those for SF-6Dv2, with an average difference of 0.080 in 77.1% of observations. At the 6-month follow-up, as shown in Fig 2c, more than 70.2% of the EQ-5D-5L utility scores outperformed the SF-6Dv2 utility scores, with a mean difference of 0.061. The limits of the agreement ranged from –0.208 to 0.330.

**Fig 2.**
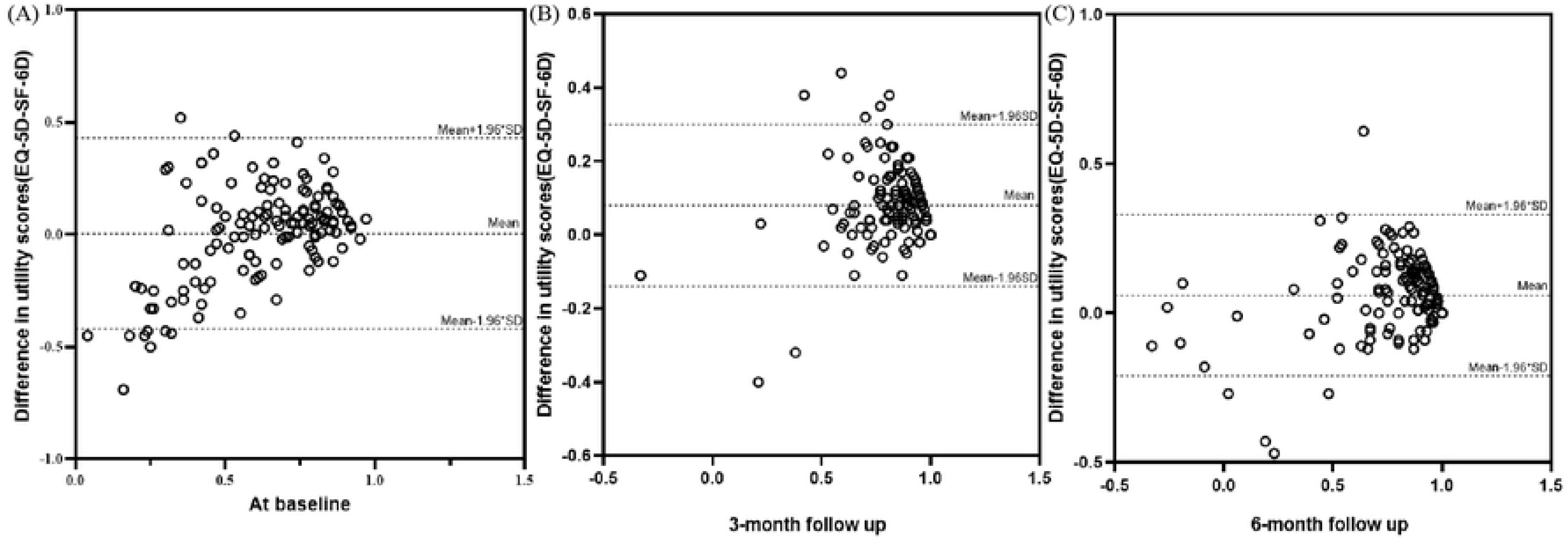
Bland–Altman plots of the difference in utility values of EQ-5D-5L and SF-6Dv2. (A) At baseline; (B) at the 3-month follow-up; and (C) at the 6-month follow-up. EQ-5D-5L: 5-level EuroQol 5-dimension. SF-6Dv2: Short Form Six-Dimension (version 2) A mean difference at the baseline of 0.082 was found between SF-6Dv2 and EQ-5D- 5L, which is lower than the recommended MID of 0.041 for SF-6D and 0.074 for EQ-5D. Fig 2c shows a mean difference of 0.080 between SF-6Dv2 and EQ-5D-5L, similar to the MID of EQ-5D but greater than the MID of SF-6D. The result of SF-6Dv2 versus EQ-5D-5L was 0.061, which was lower than the MID of EQ-5D; however, it was greater than the MID of SF-6D. It is possible that these instruments cannot be interchanged because of their clinical significance.

### Convergent validity

The correlations between the SF-6Dv2 and EQ-5D-5L dimensions based on the self-reported health of the patients are presented in Table 3. Except for the dimension of self-care (EQ-5D-5L), most correlations between SF-6Dv2 and EQ-5D-5L at baseline were moderate to weak. Indeed, the highest correlation at baseline and 3-month follow-up between the Anxiety/Depression (EQ-5D-5L) and Mental Health (SF-6Dv2) dimensions was 0.86 and 0.85, followed closely by the Role Limitation (SF-6Dv2) and Usual Activities (EQ-5D-5L) dimensions. Stronger correlations were observed at the 6-month follow-up, at which point convergent validity between similar dimensions was evident: Self-Care and Physical Function (0.41) and Self-Care and Role Limitation (0.41). The correlation for each dimension tended to be moderate to strong. Overall, the SF-6Dv2 and EQ-5D-5L utilities were highly correlated at baseline (rho = 0.709), 3-month follow-up (rho = 0.772), and 6-month follow-up (rho = 0.792).

**Table 3.**
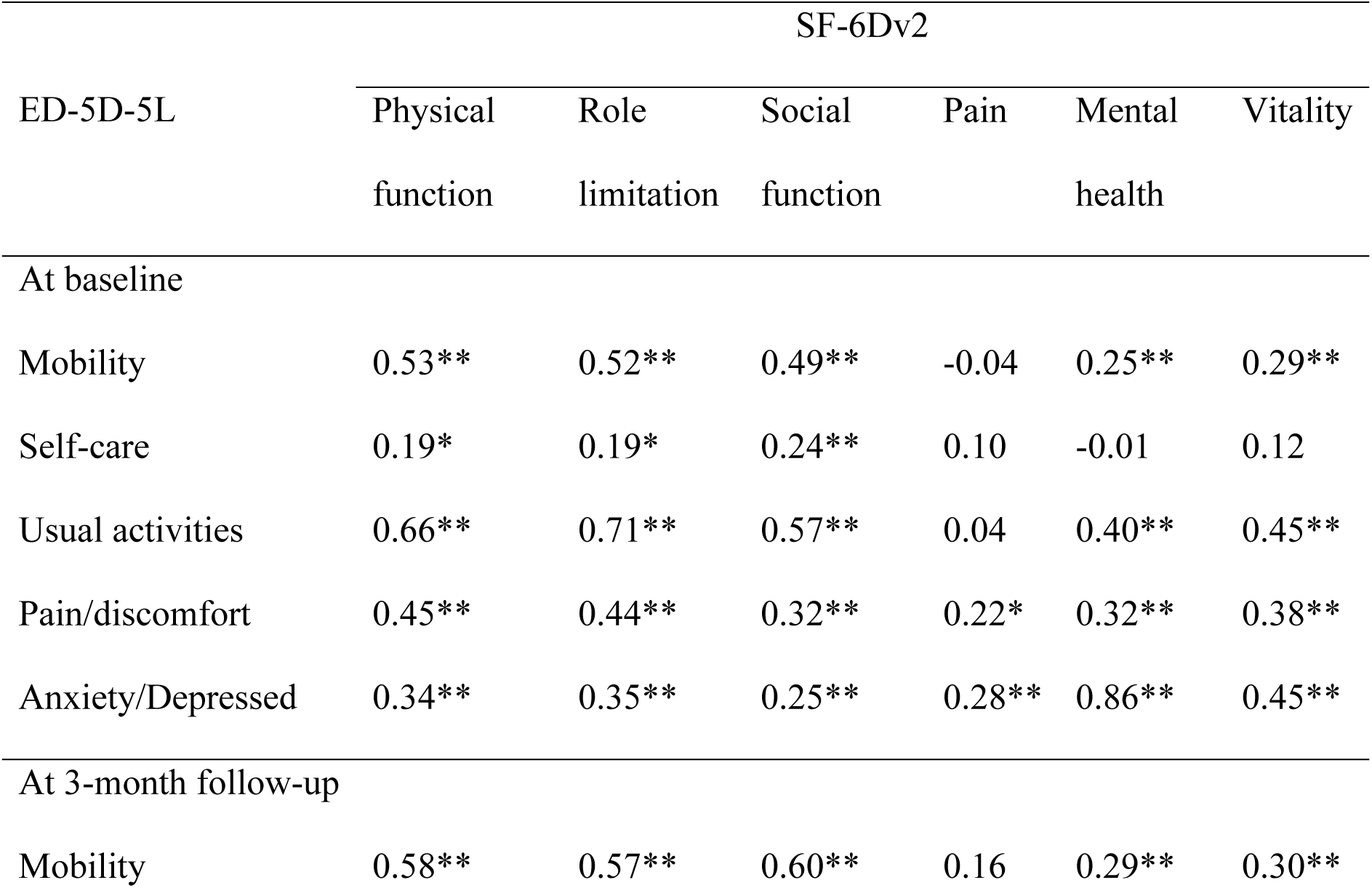
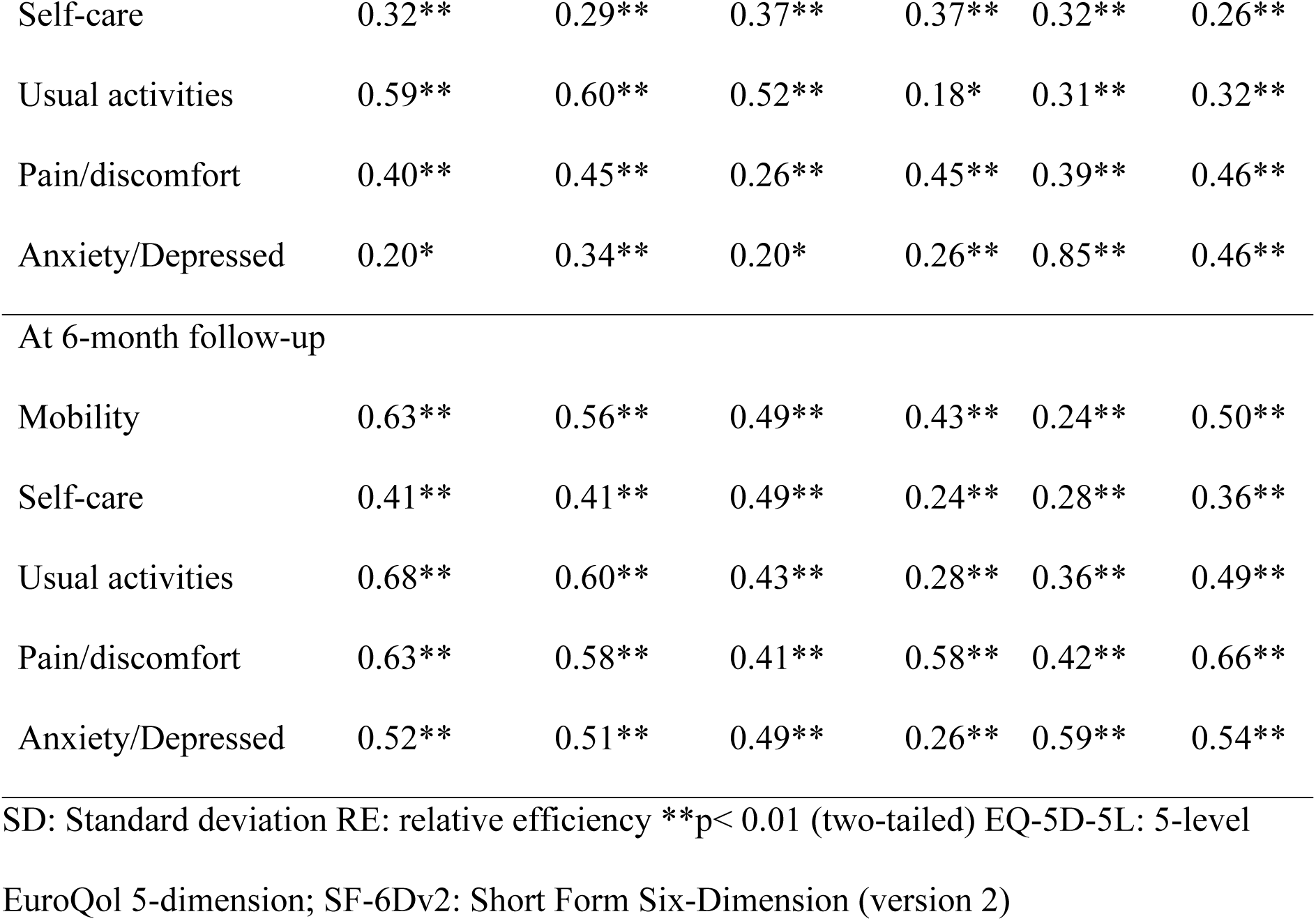
Spearman correlations among the EQ-5D-5L and SF-6Dv2 domains.

| EQ-5D-5L | SF-6Dv2 |  |  |  |  |  |
| --- | --- | --- | --- | --- | --- | --- |
|  | Physical<br>function | Role<br>limitation | Social<br>function | Pain | Mental<br>health | Vitality |
| At baseline |  |  |  |  |  |  |
| Mobility | 0.53** | 0.52** | 0.49** | -0.04 | 0.25** | 0.29** |
| Self-care | 0.19* | 0.19* | 0.24** | 0.10 | -0.01 | 0.12 |
| Usual activities | 0.66** | 0.71** | 0.57** | 0.04 | 0.40** | 0.45** |
| Pain/discomfort | 0.45** | 0.44** | 0.32** | 0.22* | 0.32** | 0.38** |
| Anxiety/Depressed | 0.34** | 0.35** | 0.25** | 0.28** | 0.86** | 0.45** |
| At 3-month follow-up |  |  |  |  |  |  |
| Mobility | 0.58** | 0.57** | 0.60** | 0.16 | 0.29** | 0.30** |
| Self-care | 0.32** | 0.29** | 0.37** | 0.37** | 0.32** | 0.26** |
| Usual activities | 0.59** | 0.60** | 0.52** | 0.18* | 0.31** | 0.32** |
| Pain/discomfort | 0.40** | 0.45** | 0.26** | 0.45** | 0.39** | 0.46** |
| Anxiety/Depressed | 0.20* | 0.34** | 0.20* | 0.26** | 0.85** | 0.46** |
| At 6-month follow-up |  |  |  |  |  |  |
| Mobility | 0.63** | 0.56** | 0.49** | 0.43** | 0.24** | 0.50** |
| Self-care | 0.41** | 0.41** | 0.49** | 0.24** | 0.28** | 0.36** |
| Usual activities | 0.68** | 0.60** | 0.43** | 0.28** | 0.36** | 0.49** |
| Pain/discomfort | 0.63** | 0.58** | 0.41** | 0.58** | 0.42** | 0.66** |
| Anxiety/Depressed | 0.52** | 0.51** | 0.49** | 0.26** | 0.59** | 0.54** |

SD: Standard deviation RE: relative efficiency \*\*p< 0.01 (two-tailed) EQ-5D-5L: 5-level
EuroQol 5-dimension; SF-6Dv2: Short Form Six-Dimension (version 2)

### External validity

SF-6Dv2 and EQ-5D-5L can distinguish between known group validity (Table 2). As measured by the SF-6Dv2 and EQ-5D-5L, participants who were less educated, unmarried, retired, or unemployed, had comorbidities ≤ 2, and higher NYHA classification tended to have lower average utility. Additionally, EQ-5D-5L was considerably more efficient concerning NYHA functional class (RE = 1.23) and current smoking (RE = 1.39). At the same time, SF-6Dv2 performed better concerning sex (RE = 0.01), annual household income (RE = 0.77), employment status (RE = 0.34), education (RE = 0.03), marital status (RE = 0.76), and comorbidities (RE = 0.22).

## Discussion

This study compared the changes over time between the generic health status questionnaires SF-6Dv2 and EQ-5D-5L in patients with HOCM. To the best of our knowledge, this is the first study to compare EQ-5D-5L and SF-6Dv2 instruments in patients with HOCM. A 6-month follow-up allowed us to investigate the sensitivity of the two scales to changes in health status. As reported above, SF-6Dv2 and EQ-5D-5L displayed some parallels with respect to the ability to detect trends in the changes in health utility values.

However, the absolute amounts of measured utility values were not identical. The two scales had similar average utility values at baseline. However, at the subsequent follow-up visit, when the physical function was better than SF-6Dv2, EQ-5D-5L generally showed higher utility in the same patient. These results were consistent with a recent study comparing the two instruments [23,24]. A previous study comparing EQ-5D-5L and SF-6Dv2 in breast cancer patients indicated that the utility value of EQ-5D-5L exceeded that of SF-6Dv2, which also agreed with our findings [25]. The percentage change from the baseline in the EQ-5D-5L questionnaire was higher. It is reasonable to assume that EQ-5D-5L is more responsive to change than SF-6Dv2. This might be partly attributed to the different dimensions covered by the two scales and the different utility ranges corresponding to each measure. The ceiling effect of EQ-5D-5L was evident as the NYHA cardiac function class improved after discharge. Our follow-up results showed a higher ceiling effect [26]. The disease’s morbidity level is thought to be a possible factor influencing the ceiling effect in EQ-5D [27].

Numerous previous studies have proven the ceiling effect of EQ-5D-5L [28,29]. These results suggest that an instrument with higher ceiling effects is ineffective in differentiating relatively better health conditions. However, neither EQ-5D-5L nor SF-6Dv2 showed floor effects. As an explanation, most patients received regular treatment on the day of admission accompanied by their family members, which led to improvements in the physical impairment and mental symptoms caused by the disease. In contrast, the SF-6Dv2 involved the period over the previous four weeks and presumably had a recall bias that prevented patients from accurately perceiving physical discomfort and mental impairment within the specified time of the instrument. After the patient was discharged from the hospital at the end of the intervention, due to home care and regular re-examination of chronic diseases, the assessment of the patient’s worst self-health status was further reduced. There is no consensus on a methodology for comparing the utility scores of different MAUIs. Owing to the lack of a distinct reference standard for basic preference measures assessing health outcomes, we compared the combination of indicators concerned with correlation, consistency, convergent validity, and known group validity.

The ICC between the utility scores of the two tools can be observed to be moderate to good, similar to previous findings [30,31]. Nevertheless, the agreement of the instruments tended to worsen in patients with a higher NYHA cardiac function classification and poorer disease, that is, patients with lower general health status. Similar findings have been found in other studies, with low consistency between the EQ-5D-5L and SF-6Dv2 in the lower health state and vice versa [32,33]. In addition, the Bland–Altman plots showed that the 95% levels of agreement for both SF-6Dv2 and EQ-5D-5L utilities were higher than the recommended threshold of minimal clinically important difference, implying that clinically significant differences existed between utilities; therefore, the specific tools could not be regarded as interchangeable. Thus, the findings may vary depending on the scale used.

Surprisingly, the “pain” domain at baseline was an exception (rho = 0. 22), which is in contrast to the results of other studies that concluded that the correlation was strongest in the area of pain [34,35]. An important consideration for inconsistencies is that pain is not the most prominent symptom of HOCM. As expected, the two questionnaires correlated well in similar domains with improved health status, especially six months after discharge. Hence, when participants possessed better physical function, the construct validity was supported by a high degree of convergence. In line with this, the phenomenon frequently appears in the academic literature on the topic of this study. As described below, when describing dimensions across comparable tools or representing the same concept owing to their names, there is a risk that it may lead to inaccurate conclusions [36]. When describing dimensions across instruments as similar or representing the same concept based on their names, there is a risk that it may lead to incorrect conclusions.

We validated the known group validity of both SF-6Dv2 and EQ-5D-5L. The RE data showed that the SF-6Dv2 had higher discriminatory efficiency than the EQ-5D-5L version for sociodemographic characteristics other than the NYHA classification and whether they are currently smoking. SF-6Dv2 displayed better construct validity than EQ-5D-5L in a similar study [9]. The reason for the greater efficiency level of SF-6Dv2 in recognizing health-related external indicators may be associated with the features of a more detailed system of description, such as physical function and role limitation dimensions [37]. However, further research is required to understand these underlying reasons better.

This study fills a data gap in measuring utility values in Chinese patients with HOCM using EQ-5D-5L and SF-6Dv2. Its main advantage is the absence of missing data due to thorough follow-up, which makes the results realistic and reliable. Another advantage of this study is that the EQ-5D-5L and SF-6Dv2 value sets were calculated using the Chinese tariff, which can precisely describe the characteristics of Chinese people. These results can be used for future cost-utility and value analyses. The first limitation of this study was the limited sample size and the single-center design. However, it should be considered that hypertrophic obstructive cardiomyopathy is rare and has a relatively low prevalence. Thus, caution should be exercised when making straightforward generalizations and extrapolating the results.

Moreover, the questionnaire was completed at baseline using face-to-face interviews and self-reports at follow-up, which may have affected the final validity to a certain degree. Finally, considering that only a few health-related external indicators were examined, a more comprehensive assessment of the content of the health survey should be conducted in the future.

## Conclusions

The data from Chinese patients with HOCM suggest that the different results obtained for SF-6Dv2 and EQ-5D-5L in the context of CUA were different. Researchers should decide to compare instruments with one another depending on the severity of the problems typically encountered in each area of the disease under study. Notably, considering the distribution advantage, ceiling effect, and discriminant validity, the SF-6Dv2 is appropriate for evaluating patients with HOCM.

## Data Availability

The datasets used and/or analyzed during the current study are available from the corresponding author on reasonable request.

## Acknowledgements

The authors sincerely thank all the inpatients who participated in the questionnaire, the assistants who helped us with the questionnaire, and the people who helped us with the mapping: Chongwen Gui (Henan University of Technology), Bowen Xie (Johnson & Johnson), and Jing Liu (Novartis).

## Supporting information Caption

**S1 Table. The ICCs of EQ-5D-5L and SF-6Dv2 index scores**

